# Comparing HIV Testing Patterns Among Adolescent Girls, Young Women, and Older Women in Nigeria: A Comparative Analysis and Socioecological Correlates

**DOI:** 10.64898/2026.08.19.26360851

**Authors:** Suliat Fehintola Akinwande, Kehinde Oluwatosin Akinwande, Carmen H. Logie, Notisha Massaquoi, Osatohanwen Joanne Okungbowa

## Abstract

**Background:** HIV testing is a key entry point for prevention and treatment in Nigeria, yet age-related disparities persist. Adolescent girls and young women (AGYW) are disproportionately affected by HIV but often demonstrate lower uptake of testing services. This study examined differences in HIV self-testing awareness, use, and antenatal HIV testing between AGYW and older women, the distribution of HIV testing service uptake, and identified multilevel factors associated with HIV self-test awareness and testing among AGYW.

**Methods:** This study analyzed cross-sectional data from the 2023-2024 Nigeria Demographic and Health Survey (NDHS), including women aged 15–49 years. Descriptive statistics were used to assess differences in HIV self-testing awareness, use, antenatal HIV testing, and place of HIV testing between Adolescent Girls and Young Women (AGYW) (15–24 years) and older women (25–49 years). Among AGYW, bivariate and multivariate logistic regression were conducted to identify socioecological factors associated with HIV self-testing, awareness and use.

**Results:** A total of 14,708 AGYW and 24,342 older women were included in the analysis. The median age of AGYW was 19 years (IQR: 17–22), compared to 35 years (IQR: 25–41) among older women. Older women reported significantly higher awareness of HIV self-testing (13.1%) compared to AGYW (8.8%). Use of the HIV self-testing kit was low overall but higher among older women (2.3%) than among AGYW (1.1%). Older women reported greater uptake of antenatal HIV testing (57.9%) than AGYW (48.4%). Education, wealth, and internet use were positively associated with awareness, while AGYW aged 15–19 had significantly lower odds of awareness than AGYW women aged 20-24. Regional disparities were also observed in the awareness and use of HIV self-test kits.

**Conclusions:** Findings support that targeted, AGYW-centred strategies are needed to improve equitable access to HIV testing and achieve national and global HIV prevention goals.

## Introduction

Adolescents and young people aged 15–24 years remain a priority population in the global HIV epidemic, yet their testing behaviours and service utilization patterns are not fully understood. In 2024, an estimated 40.8 million people were living with HIV, with 1.3 million new infections worldwide (1). Adolescents and young people constitute a substantial proportion of those affected (1), particularly in sub-Saharan Africa (SSA), reflecting the region’s disproportionate burden. This is especially significant as SSA has the world’s youngest population, with nearly 60% under 25 years of age, and its youth population is projected to exceed 830 million by 2050 (2).

Adolescent girls and young women (AGYW) are disproportionately affected by HIV, with an estimated 210,000 new HIV infections globally in 2023, most occurring in SSA (3). Their heightened vulnerability is driven by biological, social, and structural factors, including gender inequality, limited educational opportunities, violence, and economic dependence (4,5). Consequently, AGYW continue to experience higher HIV incidence rates and bear a disproportionate burden of the epidemic, particularly in SSA.

Nigeria has approximately 1.9 million people living with HIV, with AGYW disproportionately affected (6). HIV prevalence among females is three times higher than among males aged 15–19 years (0.3% vs. 0.1%) and more than four times higher among those aged 20–24 years (1.3% vs. 0.3%) (6). In 2024, an estimated 48,000 new HIV infections occurred (1), with AGYW accounting for a substantial proportion. These disparities are driven by biological, social, and structural factors, including economic vulnerability, limited educational opportunities, and gendered barriers to healthcare access (7,8).

Despite this disproportionate burden, important gaps remain in understanding how their HIV testing behaviours differ from those of older women. Existing studies frequently combine women into a single category, masking age-specific differences in HIV testing uptake and service utilization that are essential for developing targeted prevention and treatment strategies.

HIV self-test (HIVST) has emerged as an effective strategy for expanding access to HIV testing, particularly among underserved populations (9). Across SSA, HIVST has increased testing uptake, reached first-time testers, and improved testing among adolescents and young people. In Nigeria, HIVST was incorporated into national guidelines in 2019 to complement facility-based testing and address barriers such as stigma, confidentiality concerns, cost, and limited access to healthcare services (10,11).

Guided by the socioecological model, HIV testing behaviours are influenced by individual (e.g., education, HIV knowledge), household (e.g., wealth), community (e.g., residence), structural (e.g., region), and policy (e.g., HIVST guidelines) factors. (12,13). The socioecological model further suggests that the influence of multilevel factors varies across population groups according to social position and life-course circumstances. AGYW and older women occupy different social, economic, and relational contexts that shape their access to HIV information, healthcare, autonomy, and structural barriers. This study therefore compares HIVST knowledge and utilization among AGYW (15–24 years) and older women (25–49 years) in Nigeria, examining the multilevel factors associated with HIVST behaviours to inform strategies that improve HIVST uptake among AGYW.

## Methods

This study analyzed cross-sectional data from the 2023–2024 Nigeria Demographic and Health Survey (NDHS), conducted by the National Population Commission in collaboration with the Federal Ministry of Health and Social Welfare (FMoHSW) (FMoHSW et al., 2024). The NDHS used a stratified two-stage cluster sampling design. First, 1,400 enumeration areas were selected using probability proportional to size across urban and rural strata. Second, 30 households were systematically selected from each cluster, targeting approximately 42,000 households. Data were collected in 1,380 clusters, with 20 clusters excluded due to security concerns.

### Study Population

The study included women aged 15–49 years who were usual residents of the sampled households or visitors who spent the night before the survey in those households. Data were collected using standardized questionnaires in accordance with the WHO format (14). Information on HIV testing (i.e., awareness and use of HIVST kits, ever tested, recent testing, and antenatal testing) and socio-demographic characteristics was collected.

#### Outcome Variables

The outcomes were awareness and use of HIVST kits, tested for HIV as part of antenatal, and place of last HIV test. Awareness and use of HIVST were coded as binary variables (yes/no) based on self-reported responses. Antenatal HIV testing was defined as testing for HIV during antenatal care. Place of last HIV test was categorized into public facilities, private facilities, faith-based facilities, NGO facilities, and home/other settings.

#### Explanatory variables

The explanatory variables were selected based on prior literature on HIV testing practices among women in SSA and the availability of variables in the NDHS dataset (15). These variables were organized across individual, household, community, and structural levels, consistent with the socioecological framework (16).

Individual-level characteristics included respondents’ age, categorized as adolescents (15–19 years), young women (20–24 years), and older women (25–49 years). Educational attainment was categorized as no education, primary, secondary, and higher. Employment status was classified as not working, agriculture, manual (skilled/unskilled), sales/services, and Professional/clerical work. Marital status was grouped as single/divorced, married, and living with a partner. Listening to the radio, television, newspaper (not at all, less than once a week and at least once a week), use of the internet (not at all, less than once a week and at least once a week, almost every day), and having multiple sexual partners in the past 12 months (yes or no) were also included.

Household-level factors included having problems accessing healthcare due to obtaining permission to seek medical care, securing money for treatment, and distance to a health facility. Each variable was coded as binary (yes or no). Household socioeconomic status was categorized into quintiles (poorest, poorer, middle, richer, and richest).

Community-level factors included place of residence (urban or rural), and the structural-level factor was geopolitical zone (North Central, North East, North West, South East, South South, and South West).

### Statistical Analysis

Descriptive analyses were conducted to summarize all explanatory variables using frequencies and proportions. The prevalence of each outcome variable (awareness and use of HIVST kits, tested for HIV as part of antenatal, and place of HIV testing) was examined between AGYW and older women. We also examined differences in the proportion of awareness and use of HIVST kits between AGYW (15-24 years) and older women (25-49 years) across the six geopolitical zones. Chi-squared and t-tests were used to assess the difference in estimates between the groups for categorical and numerical variables, respectively.

Bivariate logistic regression analyses were first conducted to identify explanatory variables associated with the outcome of awareness and use of HIVST among AGYW (15-24 years). Variables with a *p*-value <0.20 were considered for inclusion in the multivariable models. A less stringent significance level was chosen at this stage to avoid excluding potential confounders that may become important in the presence of other variables, consistent with a purposeful selection approach (17). Multivariable logistic regression models were fitted separately for awareness and use of HIVST kits to estimate adjusted odds ratios (AORs) with corresponding 95% confidence intervals. In the final multivariable models, *p*-values <0.05 were considered statistically significant.

All analyses accounted for the complex survey design of the DHS by applying sampling weights, clustering, and stratification. Multicollinearity among explanatory variables was assessed using Variance Inflation Factors (VIF), with a threshold of VIF >10 indicating potential collinearity. Data analysis was performed using Stata version 15.0.

### Ethical Approval

This study is a secondary analysis of de-identified, publicly available data from the 2023–2024 NDHS. The original survey received ethical approval from the National Health Research Ethics Committee of Nigeria and the ICF Institutional Review Board (see S1 and S2 Appendices). The NDHS datasets were accessed with permission from the DHS Program on 26 January 2024. The datasets are fully anonymized, ensuring that individual respondents cannot be identified. As this study involved analysis of de-identified secondary data obtained through established DHS procedures, separate ethical approval was not required. Data confidentiality was ensured throughout the study by using only anonymized data and adhering to the DHS Program’s data-use and confidentiality requirements.

## Results

### Socio-demographic characteristics of the study

A total of 14,708 AGYW and 24,342 older women were included in the analysis (**Table 1**). The median age of AGYW was 19 years (IQR: 17–22), compared to 35 years (IQR: 25–41) among older women. A majority of AGYW were single or divorced (63.1%), while most older women were married (82.3%). In terms of parity, 70.0% of AGYW had no children, compared to 9.2% of older women; among the latter, a substantial proportion (67.3%) had three or more children. Wealth distribution was similar; however, more older women were in the richest quintile (24.2% vs. 19.4%). AGYW were more likely to be unemployed (56.8%), while older women were mainly engaged in sales/services (48.0%). AGYW reported greater barriers to healthcare access, including permission (12.1%), finances (45.6%), and distance (24.4%).

**Table 1.**
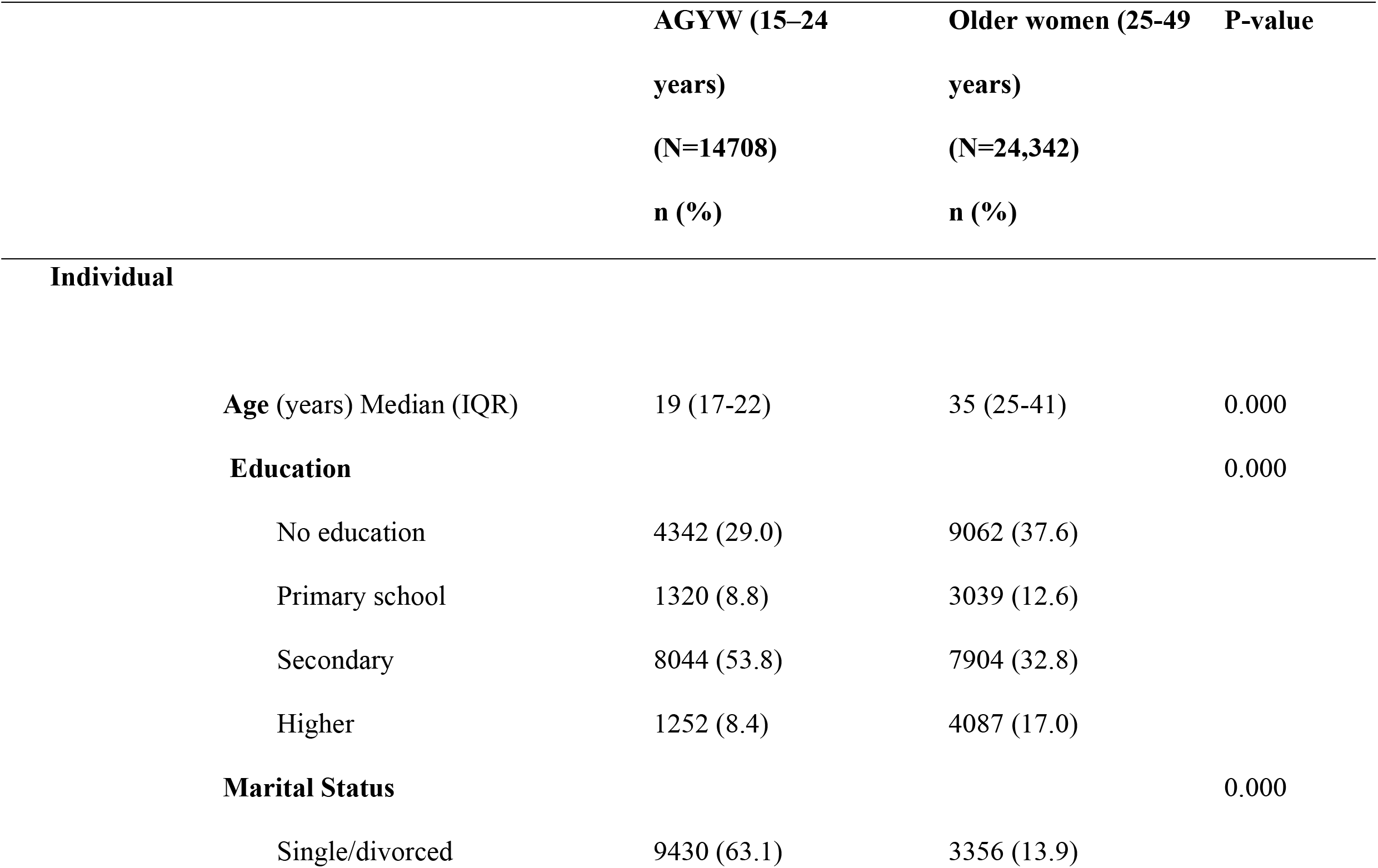
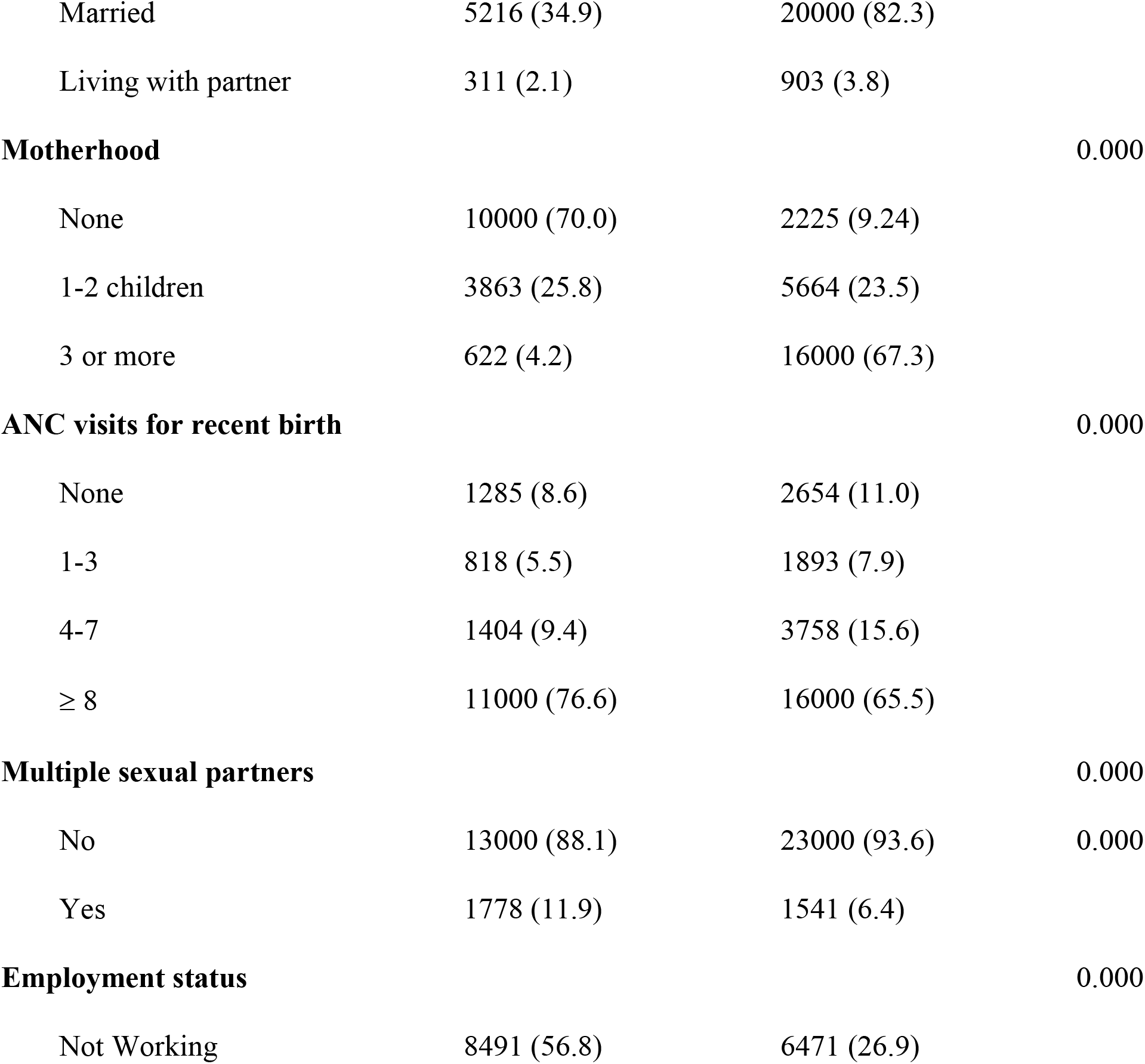
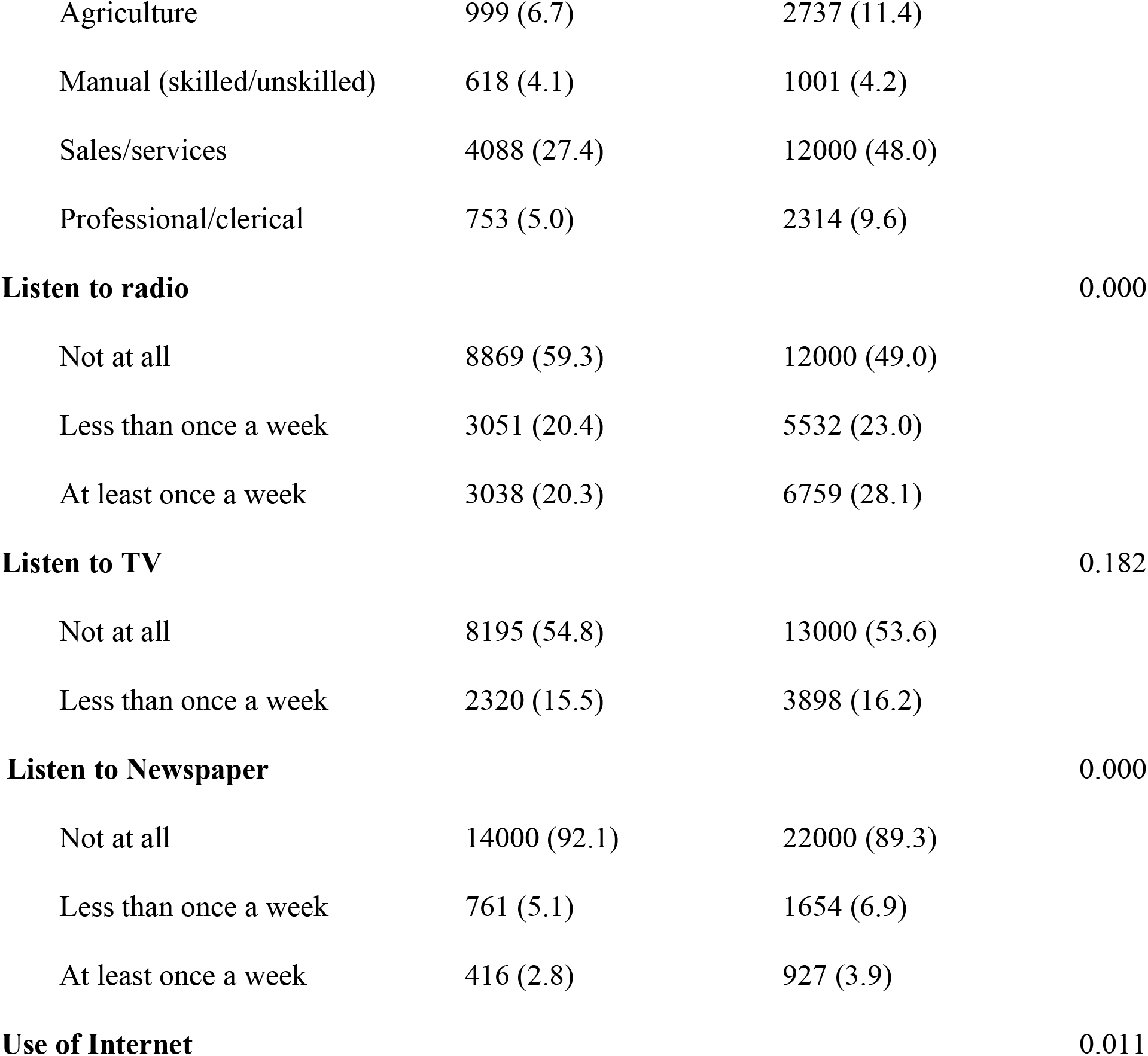
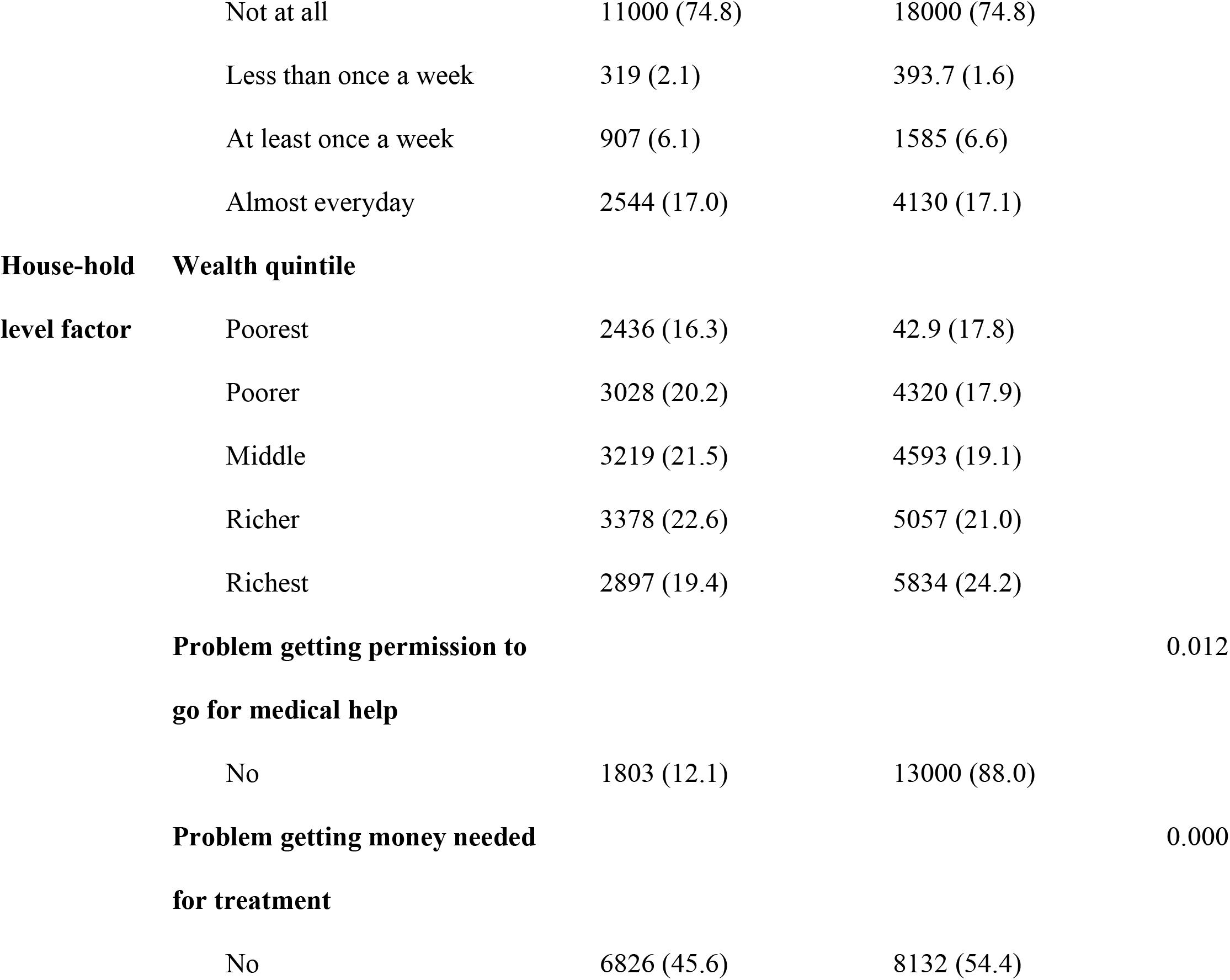
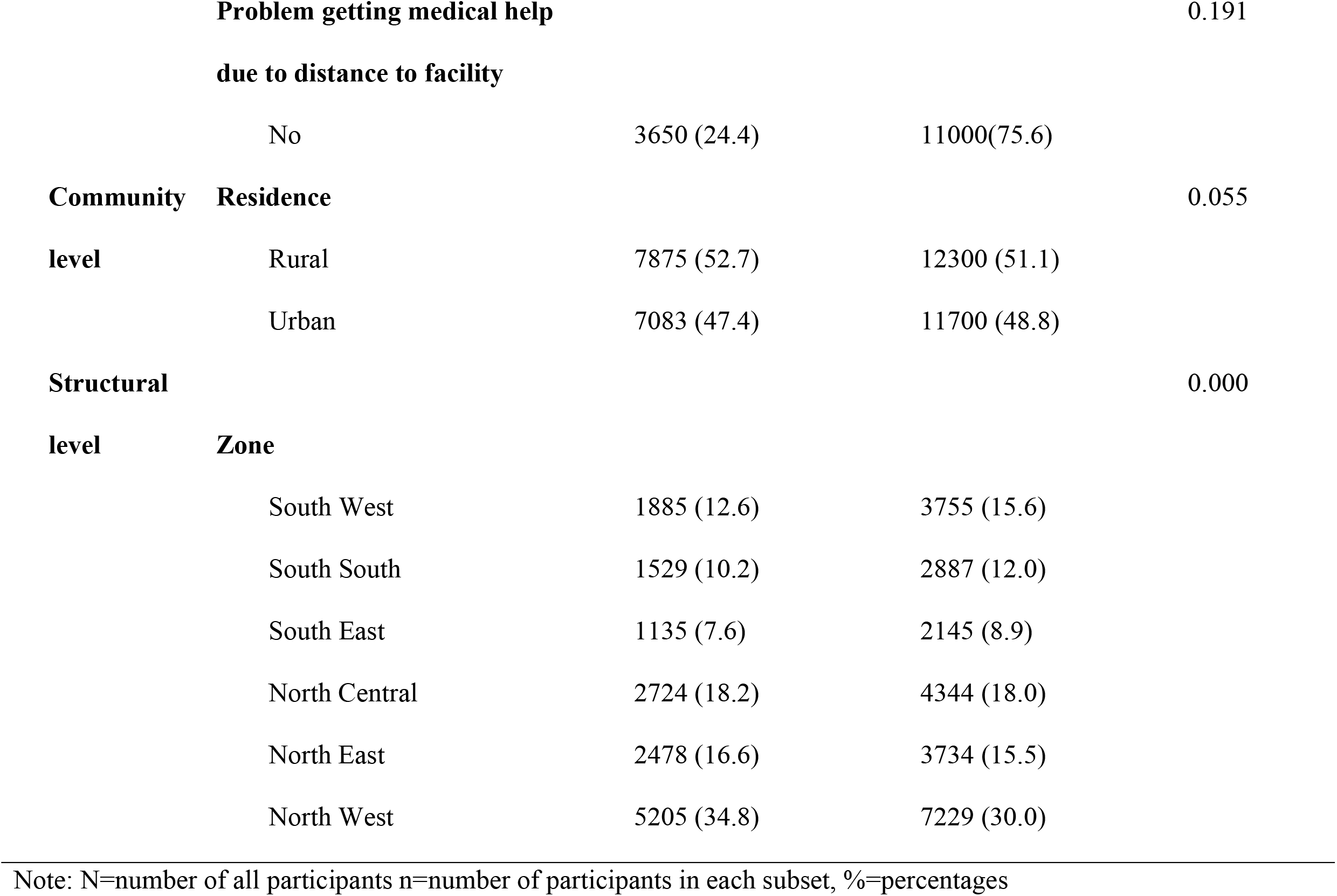
Individual, community and structural level characteristics of the study participants by AGYW and older women using NDHS data for Nigeria (2023–2024)

### Awareness and use of HIVST Kits and antenatal HIV testing in Nigeria between AGYW (15-24 years) and older women (25-49 years)

Awareness of HIVST kits was higher among older women (13.1%) than among AGYW (8.8%) (**Fig 1**). Similarly, actual use of HIVST kits remained low in both groups but was slightly higher among older women (2.3%) compared to AGYW (1.1%) (p<0.001). Antenatal HIV testing showed the highest coverage overall, with older women again reporting higher prevalence (57.9%) than AGYW (48.4%) (p<0.001).

**Fig 1.** Prevalence of awareness and use of HIV self-testing and antenatal HIV testing in Nigeria between AGYW and older women using 2023-2024 DHS Data

### Place of HIV testing

Most HIV testing occurred in the public sector for both groups, with higher use among AGYW (74.7%) compared to older women (71.2%). The private sector was the second most common testing location, also more frequently reported by AGYW (20.2%) than by older women (25.0%). Testing in faith-based facilities remained minimal in both groups but was slightly higher among older women (1.5%) than among AGYW (1.0%). Similarly, testing through NGOs or medical outreach was very low overall (0.3% among AGYW and 0.1% among older women). Home or other testing locations were less common but more frequently reported among AGYW (3.8%) compared to older women (2.3%). These differences in testing location were statistically significant (p < 0.001).

### Geographic distribution of HIVST awareness and use

Among AGYW, HIVST awareness was highest in the South South and South East zones and lowest in the North East and North West zones (Table 2). In contrast, older women reported the highest awareness in the North East and North Central zones.

**Table 2.** Awareness and use of HIV self-testing (HIVST) among AGYW (15–24 years) and older women (25–49 years) in Nigeria, 2023–2024 DHS.

| Zone | AGYW | AGYW | Older Women | Older Women |
| --- | --- | --- | --- | --- |
|  | Awareness | HIVST Use | Awareness | HIVST Use |
|  | n (%) | n (%) | n (%) | n (%) |
| South West | 187 (9.9) | 19 (1.0) | 403 (5.6) | 90 (1.2) |
| South South | 275 (18.0) | 26 (1.3) | 358 (9.6) | 74 (2.0) |
| South East | 158 (13.9) | 27 (2.4) | 724 (16.7) | 113 (2.6) |
| **North Central** | 320 (11.7) | 31 (1.1) | 420 (19.6) | 89 (4.2) |
| **North East** | 147 (5.9) | 24 (1.0) | 643 (22.3) | 103 (3.6) |
| **North West** | 231 (4.5) | 36 (0.7) | 603 (16.1) | 91 (2.4) |
| **P-value** | **<0.001** | **<0.001** | **<0.001** | **<0.001** |

Among AGYW, the highest prevalence of HIVST kits’ use was reported in the South East, while the lowest prevalence was observed in the North West (Table 2). Among older women, HIVST kits’ use was consistently higher than among AGYW across most zones. The highest prevalence was observed in the North Central and North East zones, and the lowest in the South West.

### Factors associated with HIVST awareness among AGYW (15-24 years)

#### Individual-level factors

After adjustment, adolescents aged 15–19 years had lower odds of HIVST awareness than young women aged 20–24 years (aOR = 0.68, 95% CI: 0.58–0.80) (**Table 3**). Education was associated with increased awareness: primary (aOR = 2.11, 95% CI: 1.27–3.50), secondary (aOR = 2.67, 95% CI: 1.81–3.94), and tertiary education (aOR = 4.87, 95% CI: 3.15–7.53).

**Table 3.**
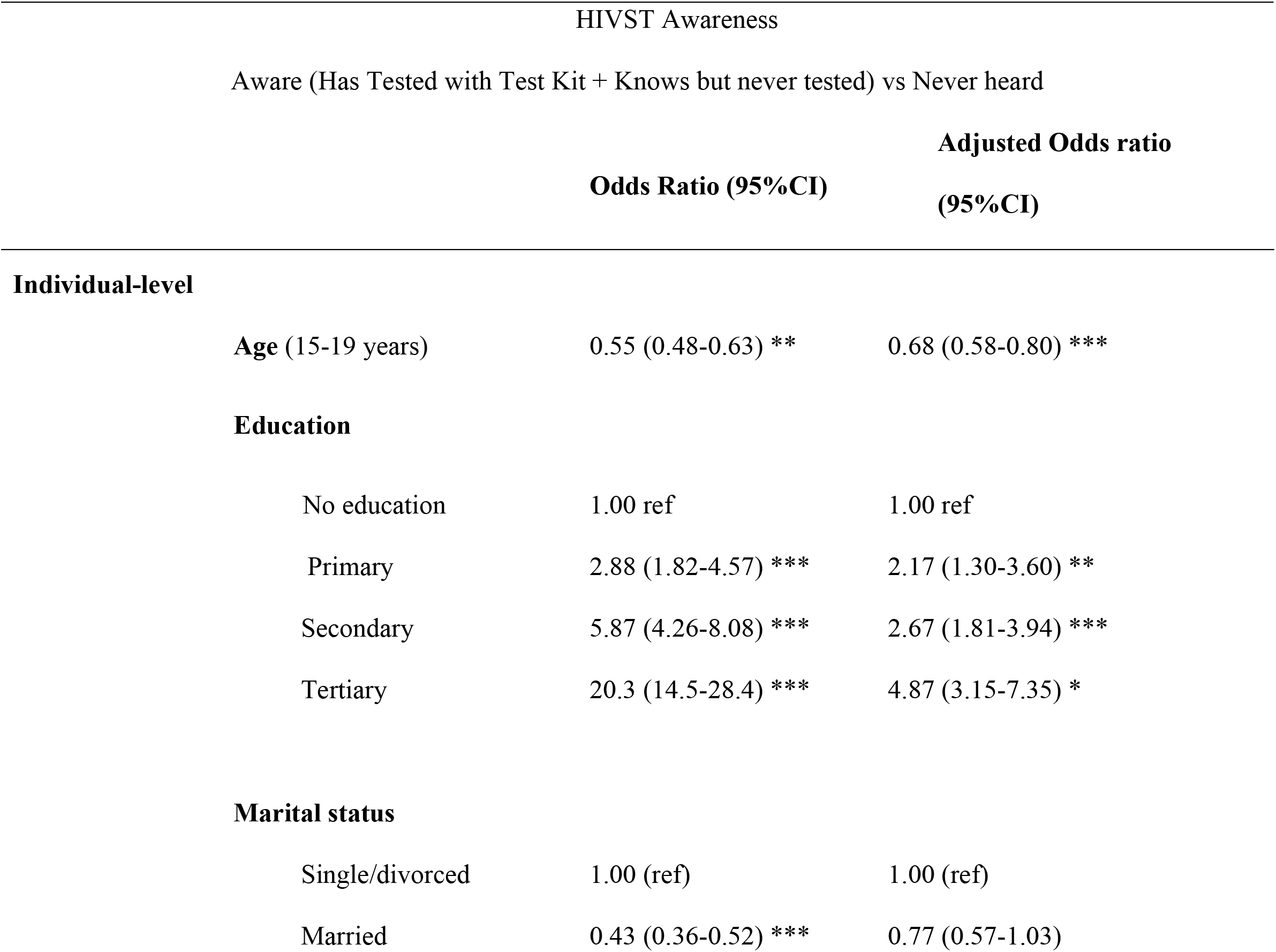
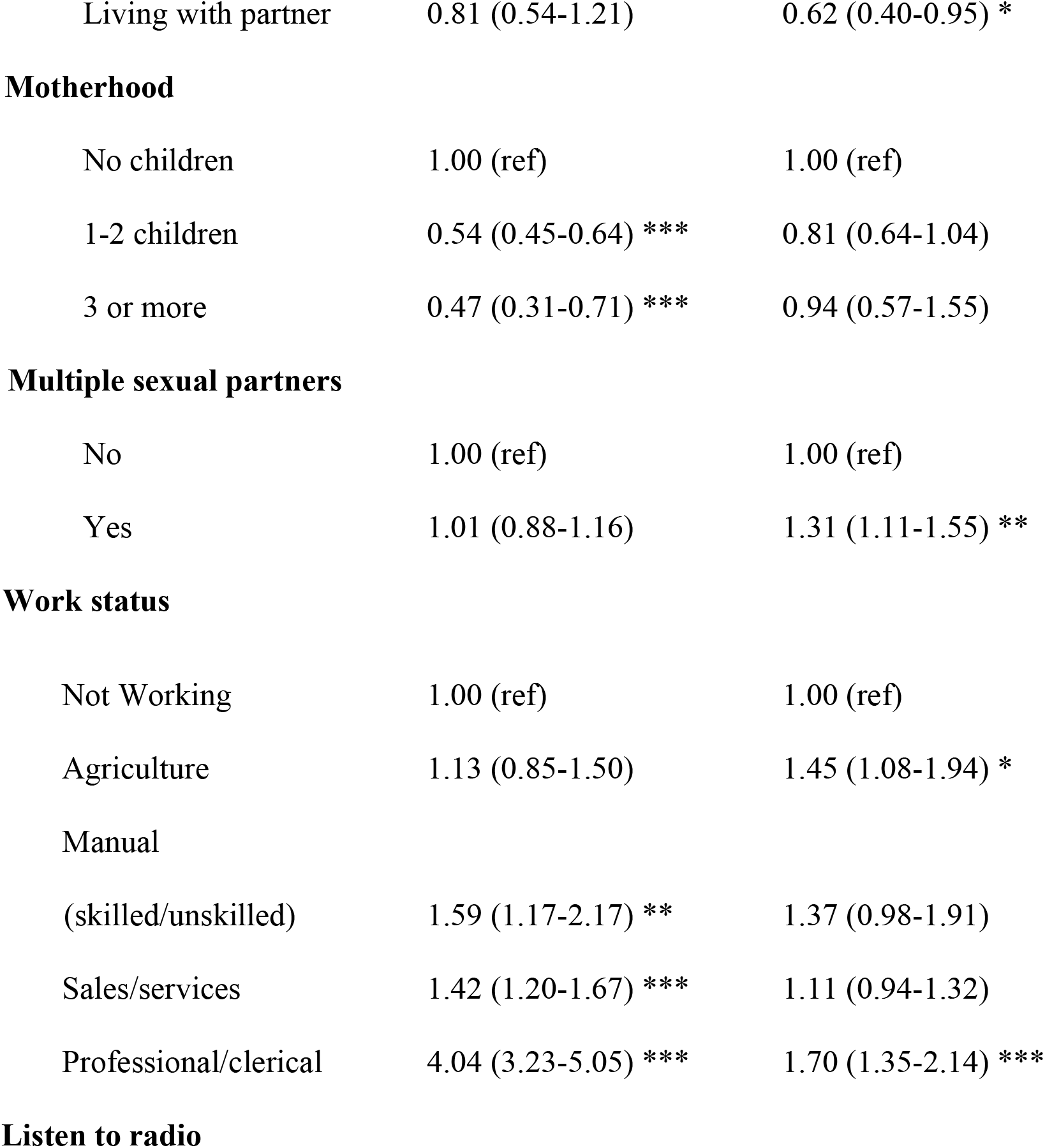
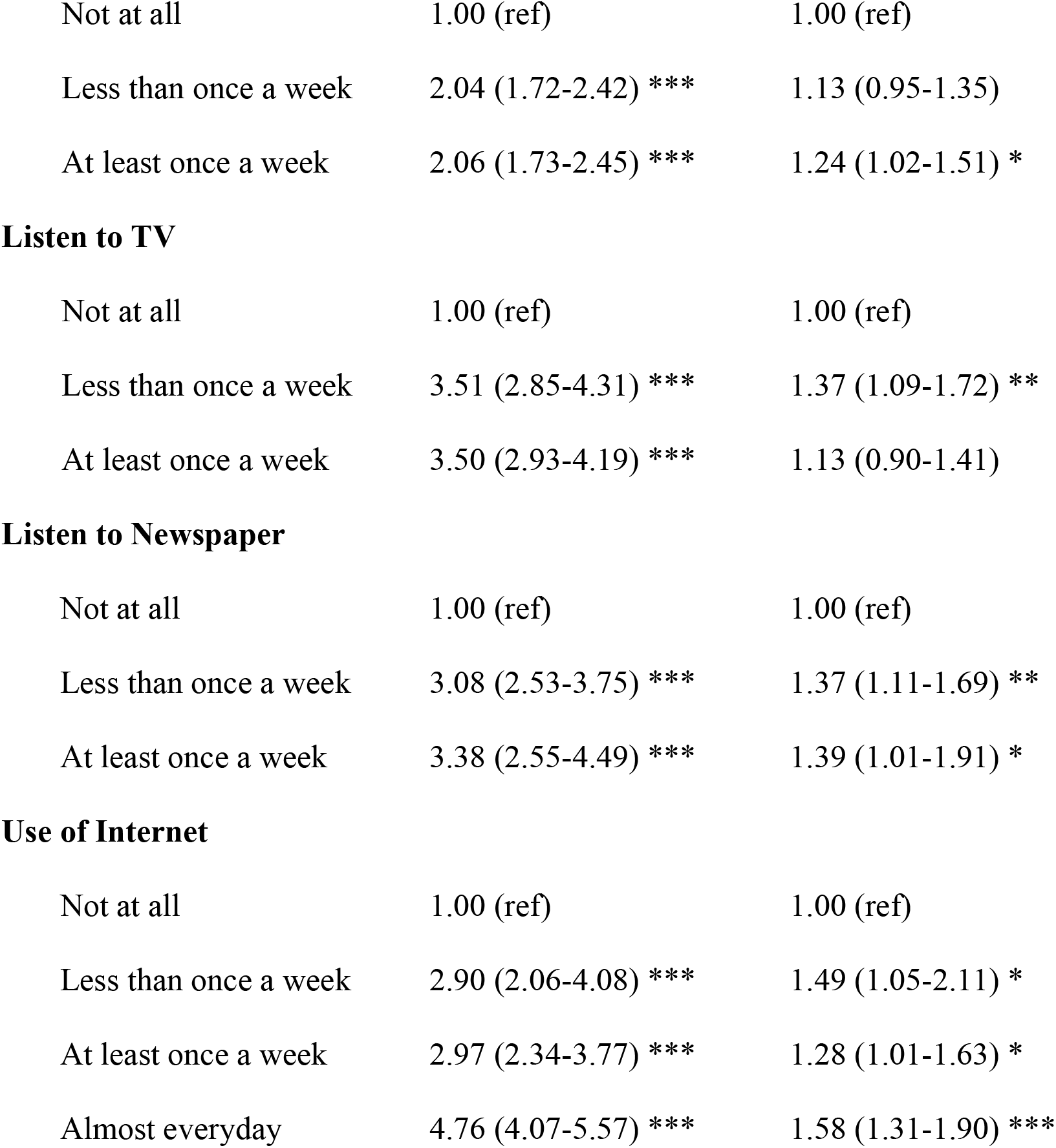
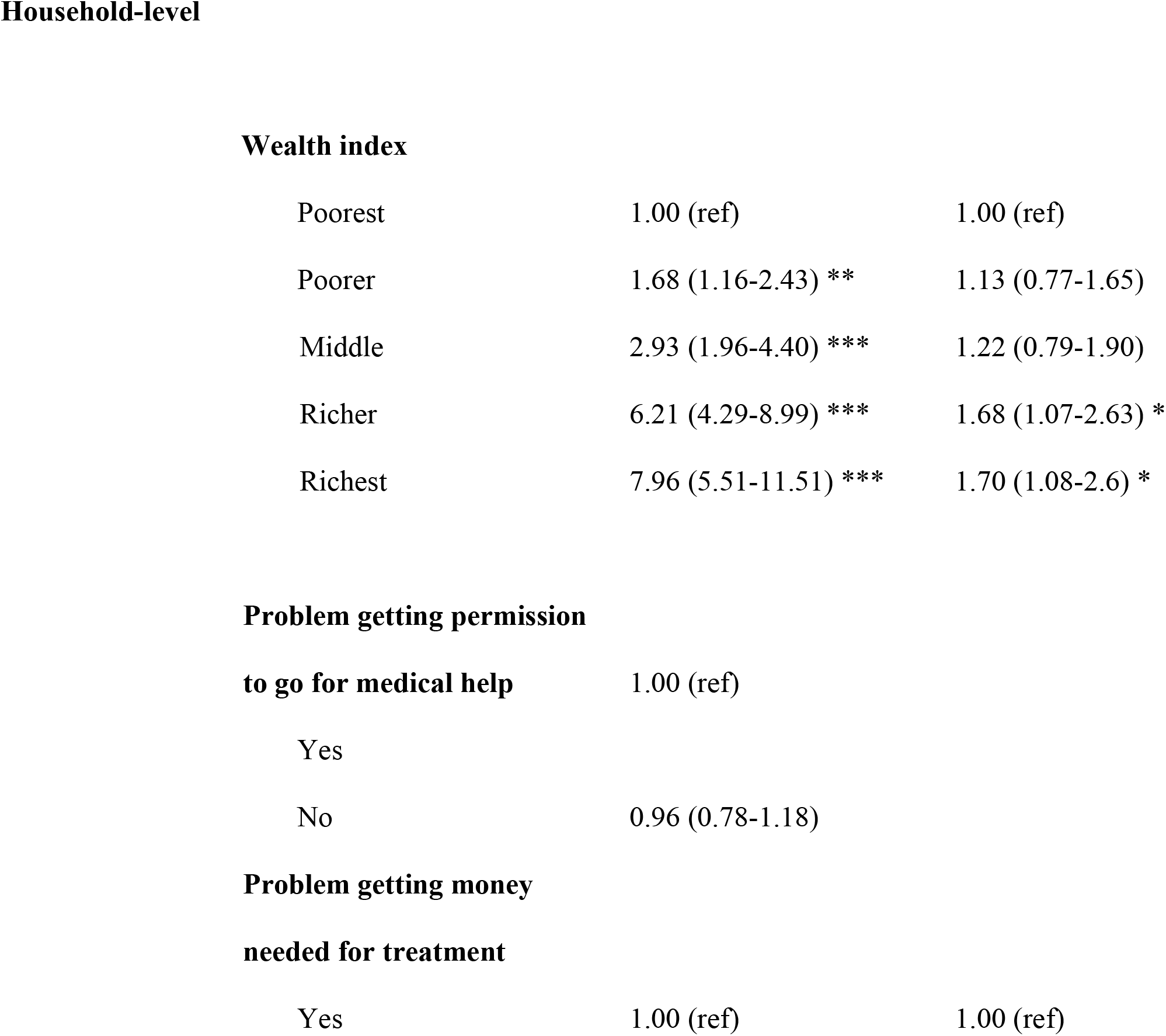
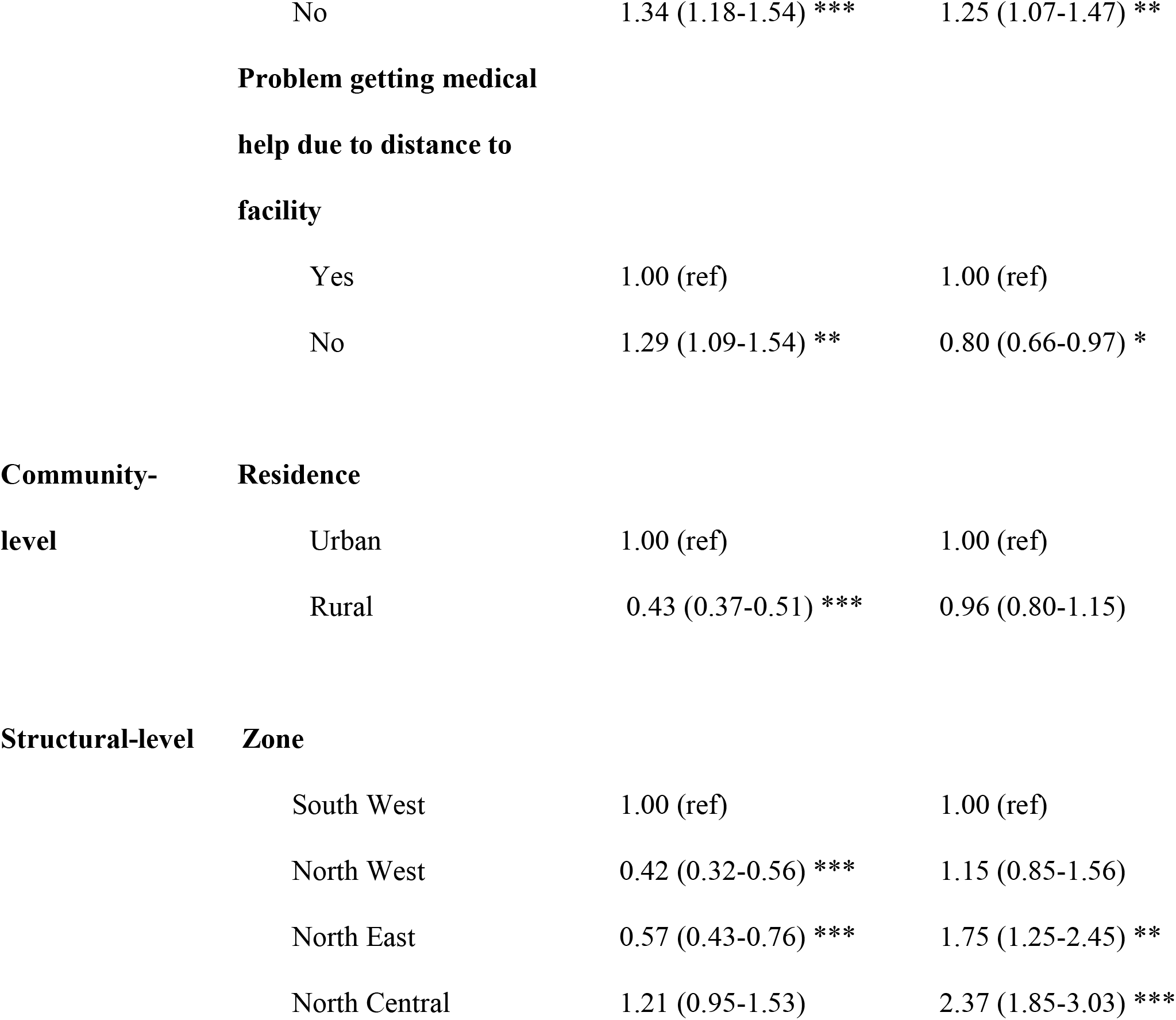
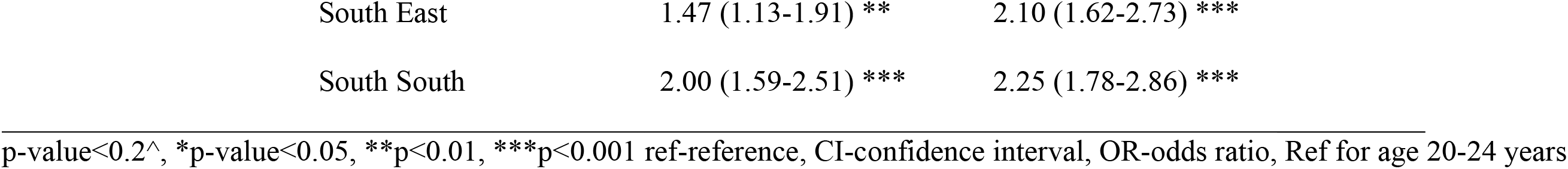
Socioecological factors associated with awareness of HIVST among AGYW in Nigeria.

Living with a partner was associated with lower awareness (aOR = 0.62, 95% CI: 0.40–0.95). Individuals with multiple sexual partners had higher odds of awareness (aOR = 1.30, 95% CI: 1.10–1.54). Employment in agriculture (aOR = 1.45, 95% CI: 1.08–1.94) and professional/clerical occupations (aOR = 1.70, 95% CI: 1.35–2.14) were associated with higher awareness. Radio, newspaper reading, and internet use were positively associated with awareness.

#### Household-level factors

Participants in the richer (aOR = 1.65, 95% CI: 1.07–2.63) and richest quintiles (aOR = 1.68, 95% CI: 1.07–2.63) had higher odds compared to the participants in the poorest.

The absence of financial barriers to healthcare was also associated with greater awareness (aOR = 1.25, 95% CI: 1.07–1.47). In contrast, reporting no distance-related barriers was associated with lower awareness (aOR = 0.80, 95% CI: 0.66–0.97).

#### Structural-level factors

Compared to the South West, higher odds of awareness were reported in the North East (aOR = 1.74, 95% CI: 1.25–2.45), North Central (aOR = 2.37, 95% CI: 1.86–3.03), South East (aOR = 2.10, 95% CI: 1.62–2.73), and South South (aOR = 2.25, 95% CI: 1.77–2.86).

### Factors associated with the use of HIVST kits among AGYW (15-24 years)

#### Individual-level factors

Younger adolescents aged 15–19 years had lower odds of HIVST compared to participants aged 20-24 (aOR = 0.54, 95% CI: 0.35–0.83). Married individuals were less likely to test (aOR = 0.38, 95% CI: 0.18–0.78) compared to single/divorced participants (**Table 4**).

**Table 4.**
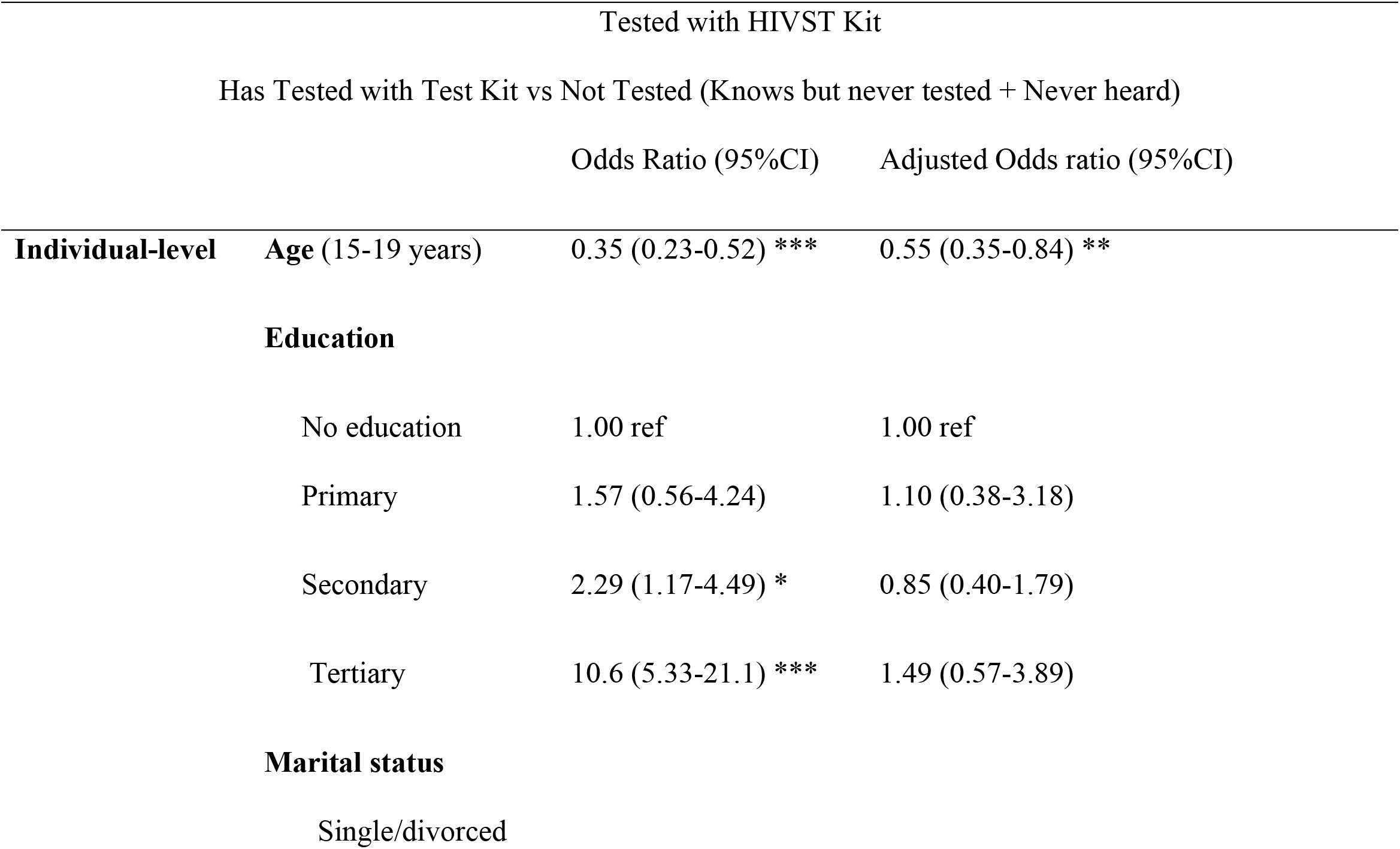
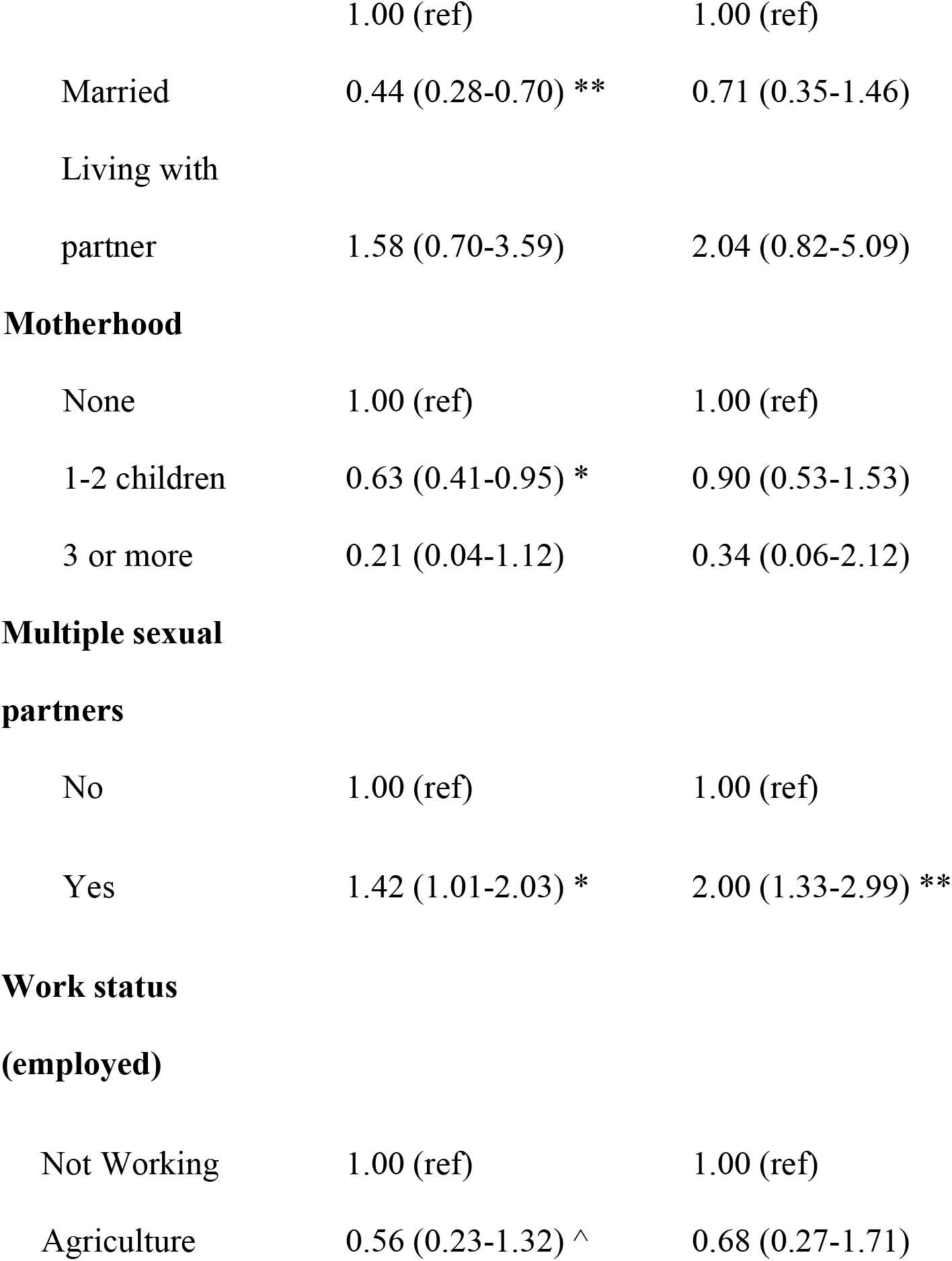
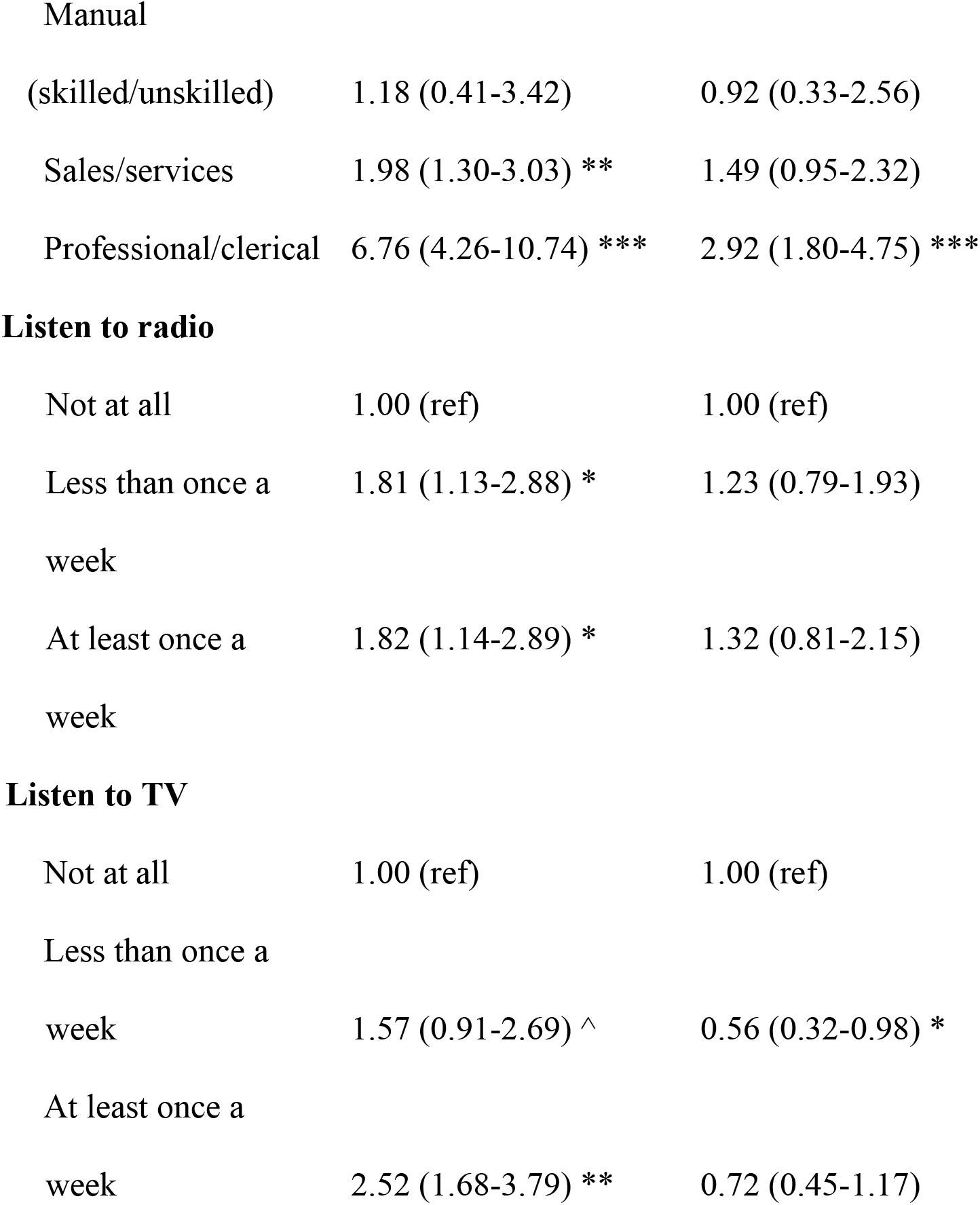
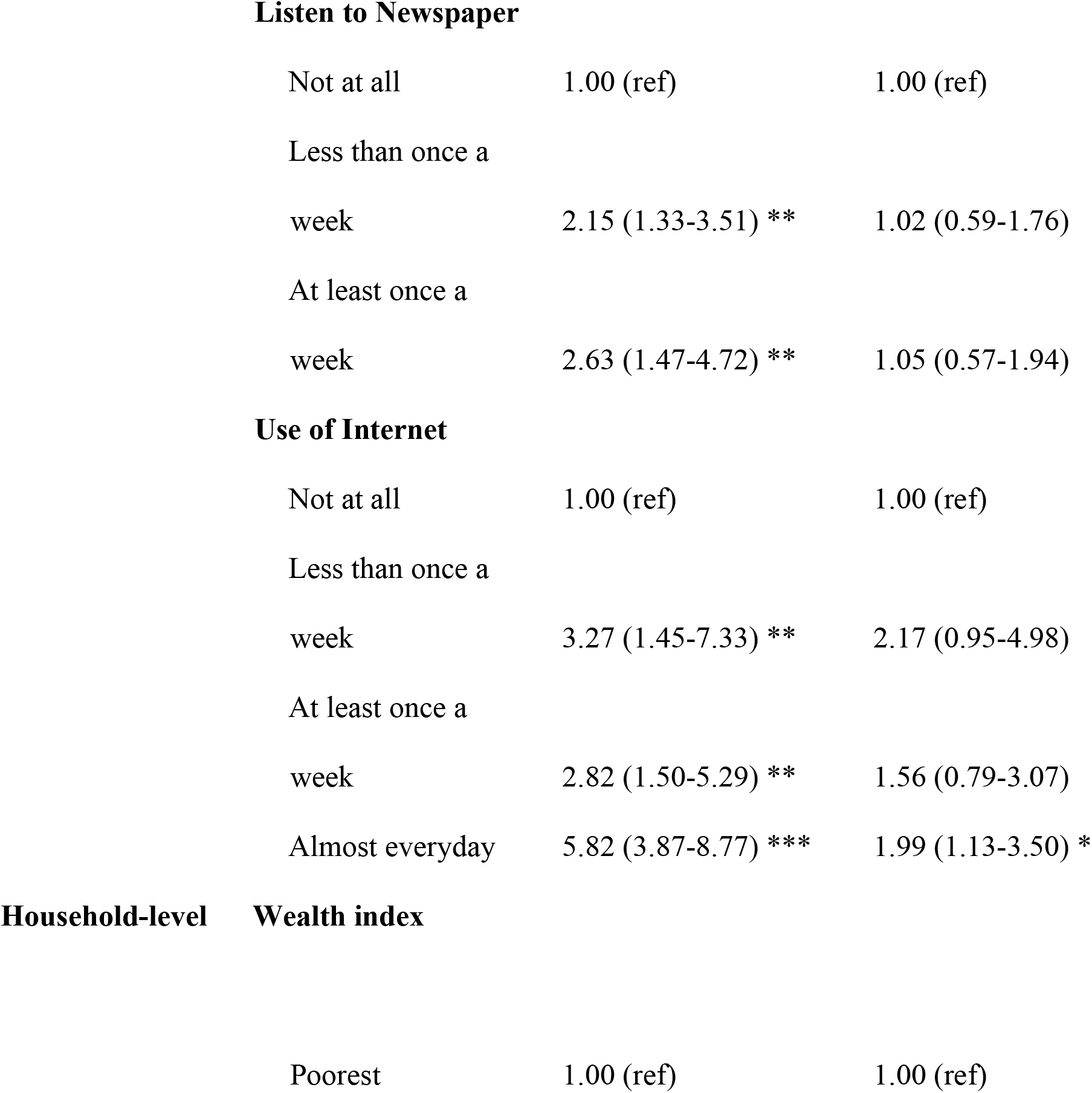
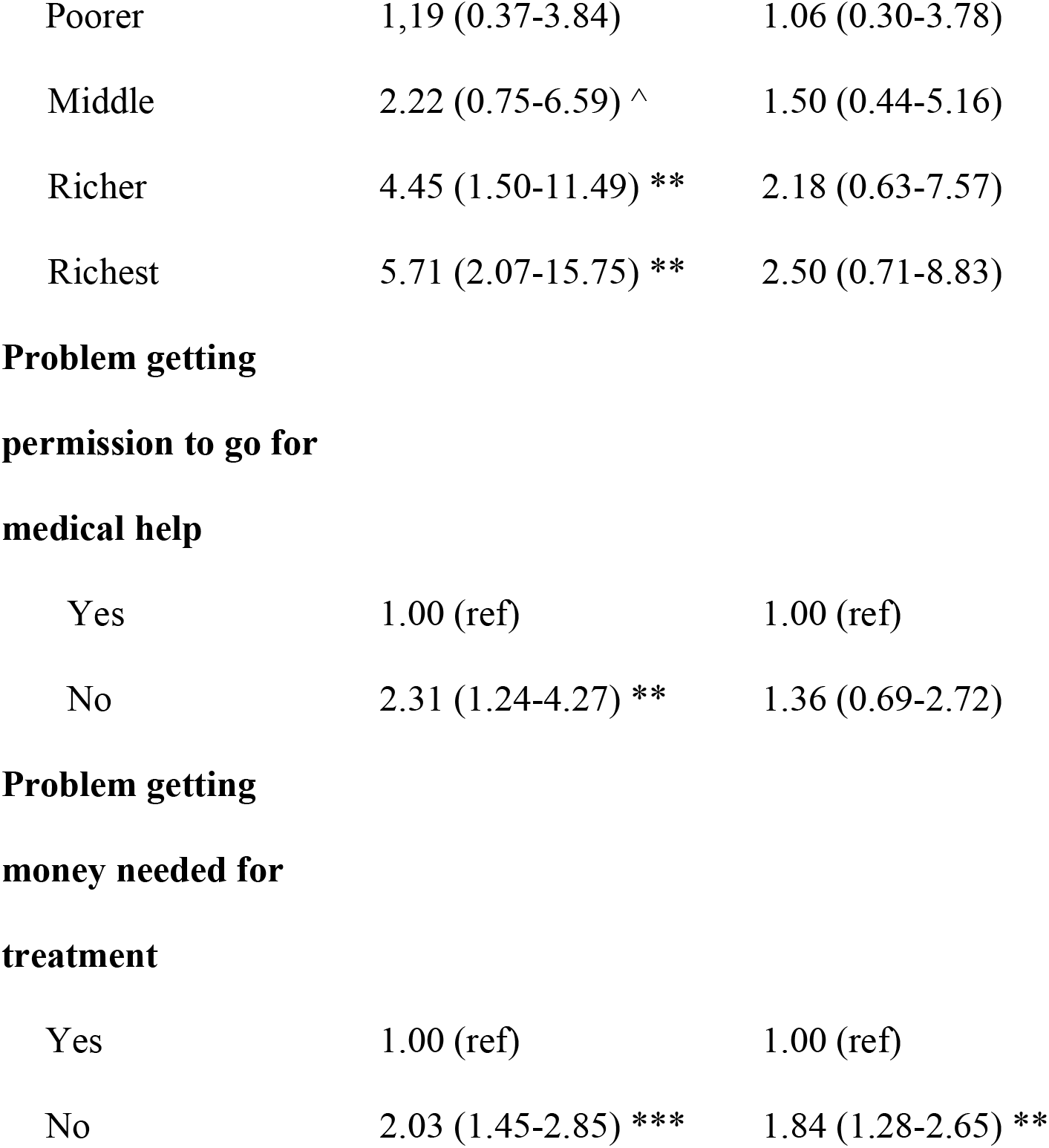
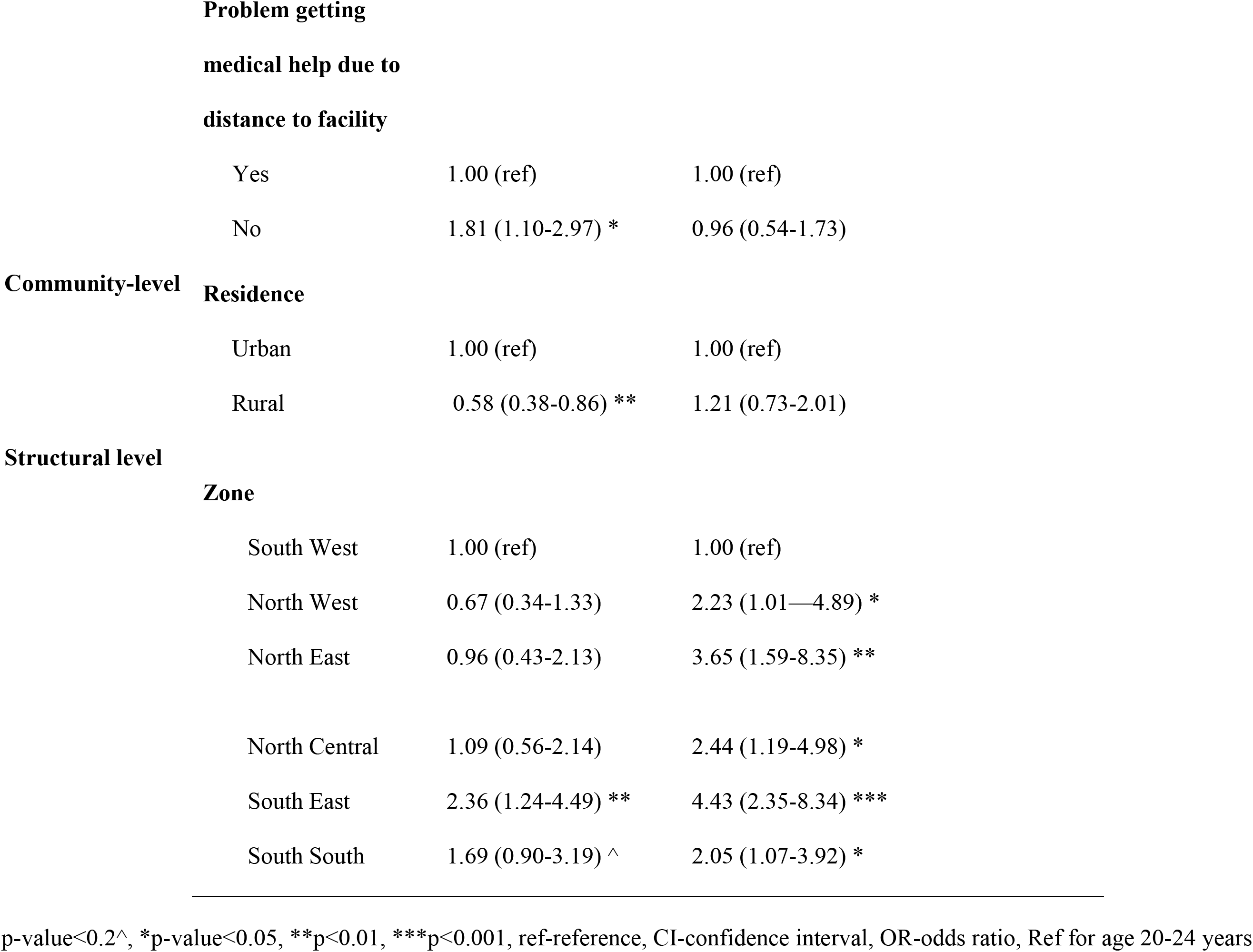
Socioecological factors associated with using the HIVST kits among AGYW in Nigeria.

Participants who had multiple sexual partners had higher odds of testing compared to those with those who did not (aOR = 1.87, 95% CI: 1.25–2.83). Employment in professional or clerical occupations was associated with increased testing (aOR = 2.94, 95% CI: 1.81–4.78). Participants who used the internet almost daily had higher odds of testing (aOR = 2.01, 95% CI: 1.15–3.52).

#### Household-level factors

Individuals who reported no financial barriers to accessing healthcare had higher odds of HIVST (aOR = 1.83, 95% CI: 1.28–2.63) compared to those who did.

#### Structural-level factors

Compared to participants in the South West, those residing in the North West (aOR = 2.23, 95% CI: 1.01–4.89), North East (aOR = 3.65, 95% CI: 1.59–8.35), North Central (aOR = 2.44, 95% CI: 1.19–4.98), South East (aOR = 4.43, 95% CI: 2.35–8.34), and South South (aOR = 2.05, 95% CI: 1.07–3.92) had higher odds of HIVST.

## Discussion

This study found that older women had higher HIVST awareness, HIVST uptake, and antenatal HIV testing than AGYW in Nigeria. HIVST awareness and use remained low overall, particularly among AGYW, highlighting persistent age-related inequities in HIV testing.

Consistent with previous studies in Nigeria and SSA, AGYW were less likely to utilize HIV testing services than older women (18). Although HIVST has been promoted as a strategy to improve testing among underserved populations, awareness and use were low, particularly among AGYW. The gap between awareness and uptake suggests that barriers, including cost, accessibility, confidentiality concerns, and limited knowledge of HIVST access pathways, continue to constrain use (19,20). Importantly, our findings reveal that awareness does not necessarily translate into use, suggesting persistent gaps in implementation and access pathways.

At the individual level, younger age and lower education were associated with lower HIVST awareness. Adolescents aged 15–19 had significantly lower odds of awareness compared to older youth, highlighting important developmental and structural barriers (21,22). Education demonstrated a clear gradient, with increasing levels associated with higher awareness, consistent with prior research linking educational attainment to improved health literacy and service utilization (18). Behavioural and informational pathways also played a role. Internet use was positively associated with awareness, suggesting that digital platforms could strengthen HIVST promotion among AGYW. However, given the persistent gaps among AGYW, these platforms may not be effectively leveraged to promote HIVST awareness and uptake among this population.

Household economic resources were positively associated with HIVST awareness and use, highlighting the influence of socioeconomic disadvantage on access to HIV prevention services (23). In contrast, distance to health facilities was not independently associated with awareness, suggesting that informational and social barriers may be more important than geographic access. Prior studies highlight the importance of informational access and social influences—such as media exposure and peer networks—in shaping HIV-related knowledge and testing uptake (24).

Significant regional disparities in HIVST awareness and use were also observed. Regional differences likely reflect variations in socioeconomic development, health infrastructure, education, and HIV programming across Nigeria’s geopolitical zones (25). Differences between AGYW and older women further suggest that regional contexts interact with age-related vulnerabilities.

Furthermore, antenatal HIV testing recorded the highest coverage among all indicators, particularly among older women, demonstrating the continued importance of maternal healthcare platforms as entry points for HIV testing. However, reliance on antenatal services may leave non-pregnant women underserved. These findings suggest that while antenatal platforms are effective, they are insufficient as a sole strategy for achieving equitable HIV testing access.

Consistent with the socioecological model, HIVST behaviours were influenced by interacting individual, household, and structural factors. Reducing disparities among AGYW will require multilevel interventions that improve affordability, expand youth-friendly distribution channels, strengthen digital engagement, and integrate HIVST into schools, pharmacies, and community programmes.

### Implication for Policy

These findings highlight the need for targeted, youth-centred interventions to improve HIVST awareness and uptake among AGYW. First, there is a need to expand access to affordable and widely distributed HIVST kits, particularly through youth-friendly platforms such as digital channels, pharmacies, and community-based programs (19,26). Secondly, leveraging digital health strategies, given the strong association between internet use and awareness, may offer a promising avenue for expanding reach among AGYW. Thirdly, integrating HIVST into non-traditional service delivery platforms, including schools and youth programs, may help bridge gaps for those outside formal healthcare systems. Importantly, these recommendations should be grounded in a gender health equity perspective, which recognizes that health policies and interventions are shaped by gendered systems of power and inequality (27).

### Study limitations

The cross-sectional design limits causal inference, and self-reported data may introduce recall and social desirability biases. NDHS data does not capture contextual nuances such as stigma and social barriers influencing HIV testing behaviours. Additionally, although the socioecological framework included policy-level constructs (e.g., national guidelines and youth-friendly policies), these factors were conceptual and not directly measured in the dataset. Despite these limitations, the study’s strengths include the use of nationally representative data and a socioecological framework, allowing for a comprehensive examination of multilevel determinants of HIV testing in Nigeria.

## Conclusion

In conclusion, this study demonstrates that awareness and use of the HIVST kit is lower among AGYW compared to older women in Nigeria. Among AGYW, these are shaped by multilevel factors spanning individual, household, and structural contexts. Addressing these disparities requires integrated strategies that move beyond traditional service delivery models to include youth-focused, context-specific, and equity-driven approaches to HIV testing and prevention.

## Data Availability

The data that support the findings of this study are publicly available from The DHS Program upon reasonable request and approval at https://dhsprogram.com

## Supplementary Document

S1 Appendix. Ethics Approval Document from National Health Research Ethics Committee

S2 Appendix. ICF IRB’s Approval

